# Hospitalization costs of patients with diabetes mellitus in Brazil, from 2013 to 2023: a time series study

**DOI:** 10.1101/2025.04.02.25325157

**Authors:** Luciana Lima de Araújo, Thaiza Teixeira Xavier Nobre, Ketyllem Tayanne da Silva Costa, Maria Alice da Silva Oliveira, Maria Eduarda Silva do Nascimento, Richardson Augusto Rosendo da Silva, Kleyton Santos de Medeiros, Clemente Neves Sousa, Aurora del Carmen Rosell Soria, Ana Elza Oliveira de Mendonça

**Affiliations:** Master in Public Health, Department of Public Health, Federal University of Rio Grande do Norte, Natal, Brazil; Doctor in Health Science. Department of Public Health, Federal University of Rio Grande do Norte, Natal, Brazil; Nurse. Department of Public Health, Federal University of Rio Grande do Norte, Natal, Brazil; Nursing Student. Department of Nursing, Federal University of Rio Grande do Norte, Natal, Brazil; Doctor in Nursing. Nursing Department, Federal University of Rio Grande do Norte, Natal, Brazil; Doctor in Health Science. Instituto de Ensino, Pesquisa e Inovação, Liga Contra o Câncer, Natal, Brazil; Doctor in Nursing. Escola Superior de Enfermagem do Porto. Porto, Portugal; Doctor of Biotechnology. University Hospital Getúlio Vargas, Federal University of Amazonas, Manaus, Brazil; Doctor in Health Science. Federal University of Rio Grande do Norte, Natal, Brazil

**Keywords:** Healthcare Financing, Hospital Costs, Diabetes Mellitus

## Abstract

**Background:** Diabetes mellitus can increase hospitalization time, requiring a multidisciplinary team and specific, high-cost supplies. In this context, the study aimed to analyze the total costs, including hospital services and professional services, of hospitalizations of patients with diabetes mellitus in Brazil between 2013 and 2023.

**Methods:** This is a time series study developed using secondary data collected from the Hospital Information System. Data collection focused on the annual costs recorded by Brazil’s Unified Health System for hospitalizations of patients with diabetes mellitus between 2013 and 2023, categorized by Brazilian states.

**Results:** When analyzing total costs in relation to each state’s Gross Domestic Product, the scenario highlights the Northeast and North regions. Hospital service costs stand out compared to professional service costs.

**Conclusions:** Diabetes mellitus requires significant investments in continuous treatments, medications, and specialized healthcare services. Beyond the financial impact, the losses for both society and individuals are considerable.

## Background

Diabetes Mellitus (DM) is among the most prevalent chronic diseases today, affecting approximately 415 million people worldwide. It is estimated that this number will reach even more alarming proportions by 2040, with an expected population of approximately 662 million people with DM [1]. A study indicated that DM is one of the most prevalent diseases among the elderly, estimating that around 18.6% of individuals aged 60 to 79 suffer from this condition. In Brazil, more than 20% of elderly individuals have DM, totaling approximately 3.5 million people [2].

DM can negatively influence the development or worsening of other comorbidities, such as cardiovascular and kidney diseases, increasing the risk of hospitalization and death. It can also extend hospitalization time, requiring a multidisciplinary team and specific, high-cost supplies. However, DM is a disease that can be prevented or controlled, reducing the risk of severe complications for patients. Therefore, adopting healthy lifestyle habits, such as engaging in physical activity, maintaining a healthy diet, and reducing alcohol and tobacco consumption, is essential [3].

Given this scenario, the study holds scientific relevance by providing a comprehensive analysis of the costs incurred in Brazil, a country with a large population of individuals with DM. This analysis can help in the development of strategies aimed at reducing hospitalization costs through investments that prioritize the prevention and control of this condition.

In this context, the study aimed to analyze the total costs, as well as the costs of hospital services and professional services, related to the hospitalization of patients with diabetes mellitus in Brazil between 2013 and 2023.

## Methods

This is an ecological study developed using secondary data collected from the Hospital Information System of SUS (SIH/SUS), a platform managed by the Department of Informatics of the Unified Health System (DATASUS). Data collection focused on the annual costs recorded by Brazil’s Unified Health System (SUS) for hospitalizations of patients with Diabetes Mellitus (DM) between 2013 and 2023, categorized by Brazilian states.

The dependent variables used were the total hospitalization costs for patients with DM, as well as the costs of hospital services and professional services, which are recorded in the Hospitalization Authorization (AIH), a document that provides the data accounted for in the SIH. In addition to raw values, the study analyzed these variables in relation to each state’s Gross Domestic Product (GDP), considering the socioeconomic specificities observed across Brazil’s vast territory. The independent variables analyzed included gender, race, type of care, states, and GDP.

Descriptive analysis was used to outline the profile of the sample in this study. The analyses were conducted using the R programming language, while graphical representations were developed using Microsoft Excel. For the spatial distribution maps of costs, the 2021 state GDP values were obtained from the Institute for Applied Economic Research (IPEA) [4].

Since states with higher GDPs may naturally present higher costs, the real difference between their expenditures could be obscured. To address this, the total cost of services for each state was divided by its respective GDP and multiplied by 1,000,000. This calculation represents the cost from 2013 to 2023 for each R$ 1,000,000.00 of GDP in each state.

As this study was conducted using secondary data from publicly accessible platforms and did not involve personal information from the studied population, approval from the Research Ethics Committee (CEP) was not required.

## Results

Figure 1 presents the spatial analysis of hospitalization costs for patients with DM across Brazilian states between 2013 and 2023. Figure 1a shows the total gross expenditure for the study period, highlighting that São Paulo had the highest investment in hospitalizations, approximately 238.4 million, followed by Minas Gerais (152 million) and Bahia (86.6 million). In general, states in the North region presented the lowest hospitalization costs for DM patients.

**Fig 1.**
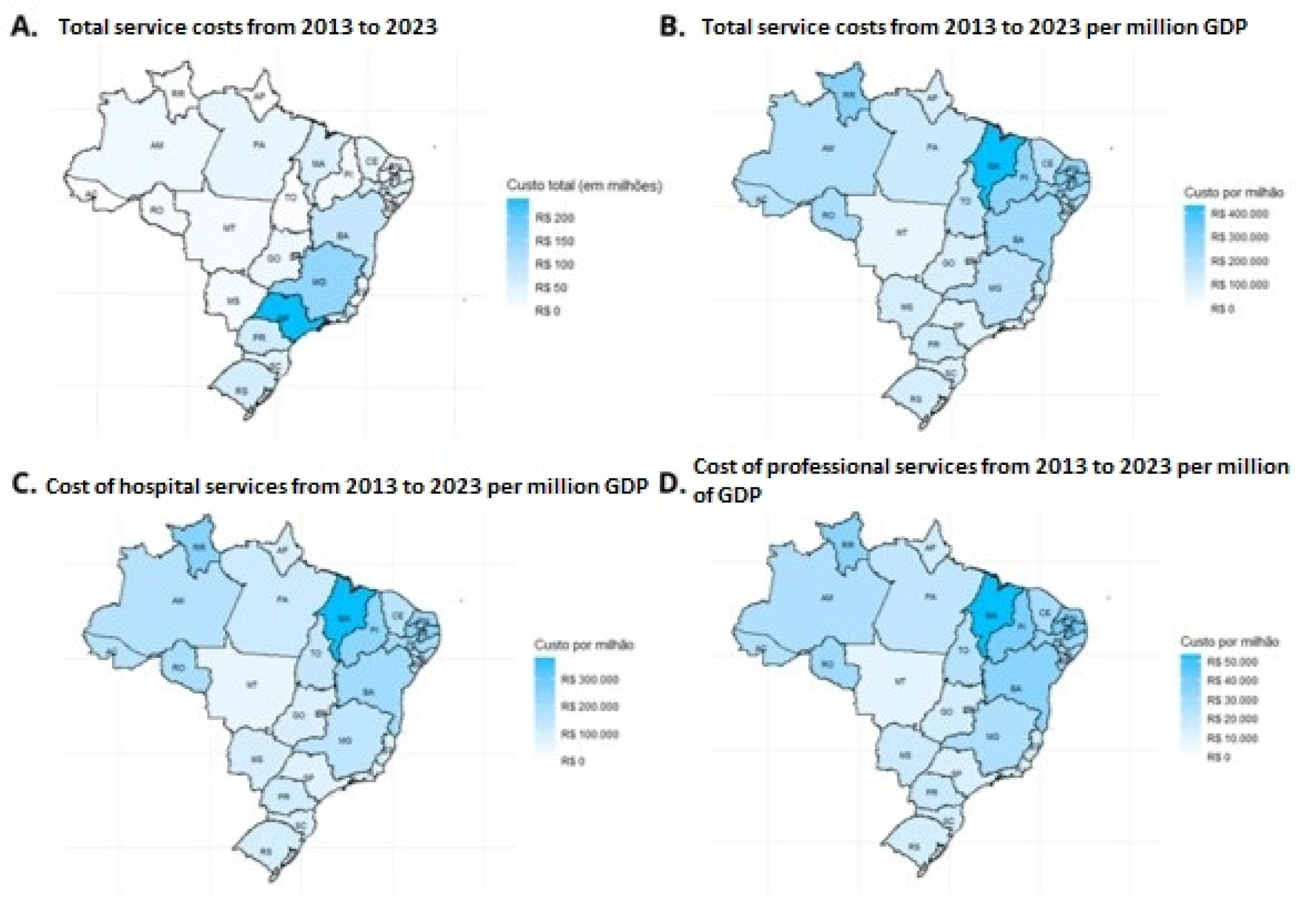
Spatial analysis of hospitalization costs for patients with Diabetes Mellitus by state in Brazil, 2013–2023. Brazil, 2024.

However, when analyzing the total costs in relation to each state’s GDP, the scenario changes, as shown in Figure 1b. In this case, the highest costs are observed in states from the Northeast and North regions, with Maranhão leading (433.57 per 1 million of GDP), followed by Paraíba (304.65 per 1 million of GDP) and Roraima (295.66 per 1 million of GDP). The lowest values are found in the Federal District (54.12 per 1 million of GDP), Mato Grosso (65.41 per 1 million of GDP), and Rio de Janeiro (75.34 per 1 million of GDP), respectively. Figures 1c and 1d further detail the costs of hospital and professional services, respectively, in relation to each state’s GDP, showing a strong resemblance to the overall scenario described in Figure 1b.

Figure 2 presents graphs analyzing costs by region. In general, total hospitalization costs (Figure 2a) have been gradually increasing, with a sharp upward trend starting in 2016. However, both the Southeast and Northeast regions show a decrease of approximately 5 million from 2022 to 2023. Meanwhile, the South region continues to rise, while the North and Central-West regions show a stabilization in their curves. As indicated in Figures 2b and 2c, hospital service costs outweigh professional service costs, with approximately 80 to 85% of total costs related to hospital services.

**Fig 2.**
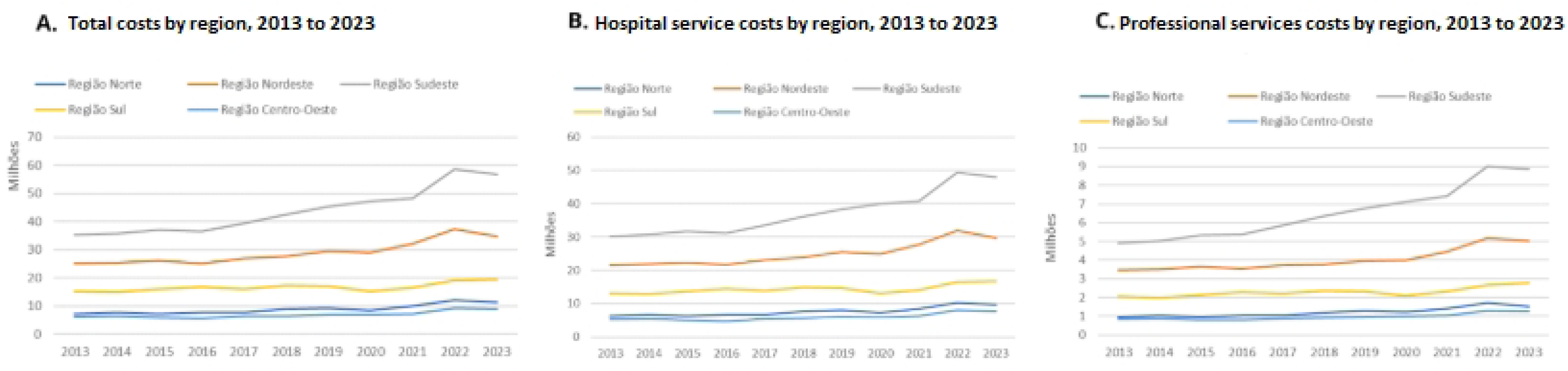
Analysis of hospitalization costs for patients with Diabetes Mellitus by region in Brazil, 2013–2023. Brazil, 2024.

Finally, Figure 3 provides an analysis of the sociodemographic profile of hospitalized DM patients between 2013 and 2023. Regarding gender, the graph indicates a higher total cost for female patients until 2015 (Figure 3a). From 2016 to 2017, the costs for both sexes equalized, but from 2018 onward, male patients incurred higher costs, mainly due to an increase in professional service expenses for this group (Figure 3c).

**Fig 3.**
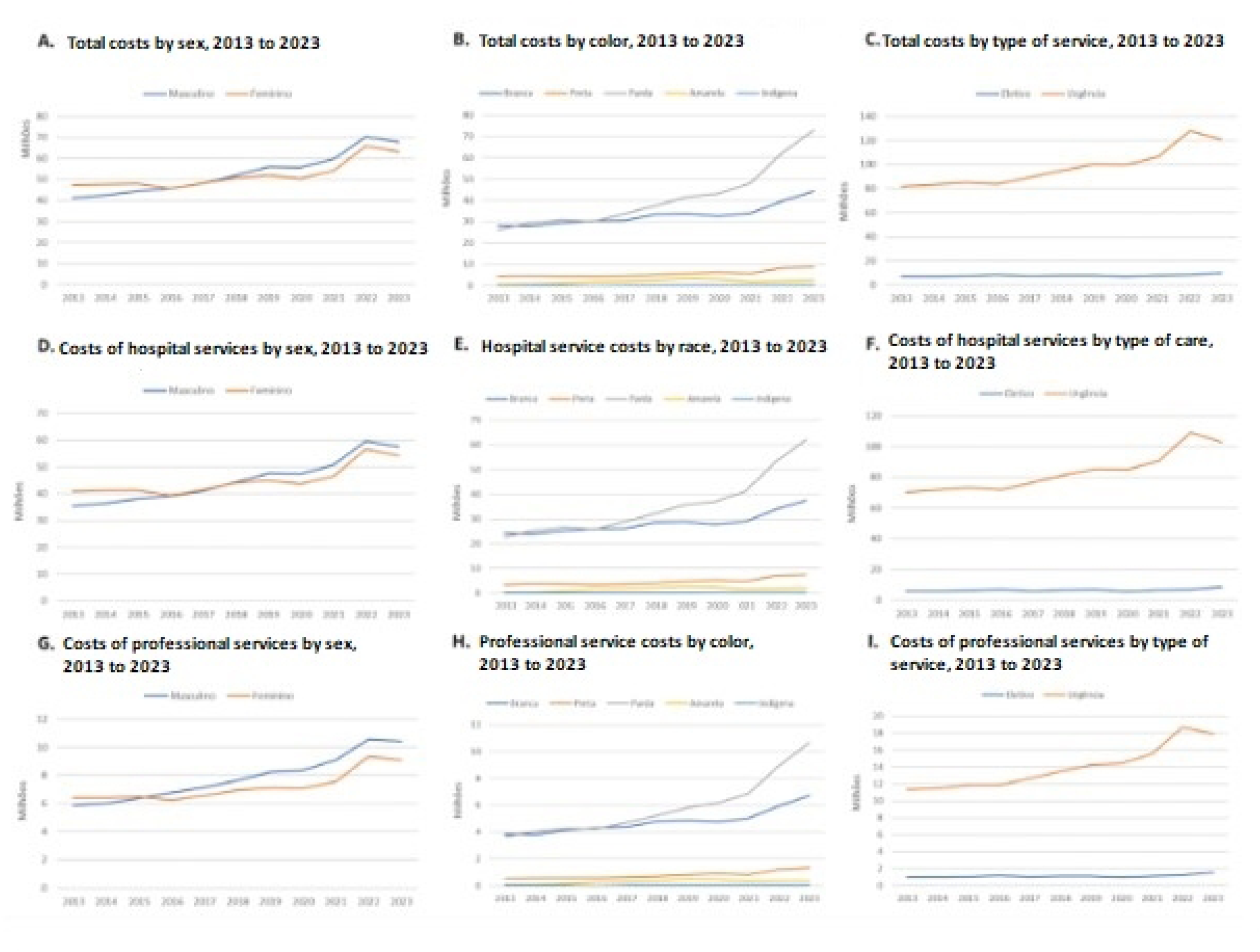
Analysis of hospitalization costs for patients with Diabetes Mellitus in Brazil by sociodemographic profile, 2013–2023. Brazil, 2024.

Regarding race, statistical analysis shown in Figures 3d, 3e, and 3f reveals that mixed-race and white patients had similar hospitalization cost indicators until 2016, when the cost for mixed-race patients saw a significant increase. This sharp rise widened the statistical gap between both racial groups, with an even more pronounced increase from 2021 onward. Figures 3g, 3h, and 3i highlight that most hospitalization costs are related to emergency care, accounting for approximately 90% of total costs.

## Discussion

In compliance with the data obtained in the study, and considering its complexity, Non-Communicable Diseases (NCDs) have a significant impact, leading to a decline in people’s quality of life, resulting in disability and a high degree of limitation in daily activities. Additionally, they are responsible for a considerable increase in mortality rates among individuals under the age of 70. This is due to systemic causes and multifactorial clinical conditions that lead to changes in the function and structure of target organs, combined with metabolic alterations that induce negative outcomes when associated with other pathologies [5].

Diabetes Mellitus primarily affects circulation, directly impairing renal function, increasing the likelihood of poor prognosis, and prolonging hospital stays. Consequently, these patients require a broader team of professionals to assist with rehabilitation and intensive care, along with specialized supplies and materials, which exponentially increase healthcare expenses [6].

Systemic alterations and chronic complications, caused by glycemic disorders and elevated glucose levels, favor the development of Diabetic Neuropathies. One of the most critical consequences of this condition is peripheral nerve involvement, which significantly limits tactile and pain sensitivity. As a result, patients are more susceptible to trauma and potential wound formation in areas where sensitivity is compromised. This, combined with tissue hypoperfusion due to circulatory impairment, creates a favorable scenario for complications arising from Diabetes Mellitus, such as delayed wound healing, amputations, and even death [7].

In cases of diabetic wounds, delayed healing is primarily caused by tissue ischemia resulting from compromised perfusion, leading to inefficient blood supply [8]. Other factors contributing to unfavorable patient outcomes include poor glycemic control, pre-existing cardiovascular diseases, ulcer formation in the lower limbs, and other peripheral arterial diseases [9].

A study conducted in all hospitals providing care under the Brazilian Unified Health System (SUS) in the state of Sergipe demonstrated that, among 109 patients hospitalized with a diagnosis of diabetic foot, 95 (87.2%) were discharged, while 14 (12.8%) progressed to death. The treatment provided ranged from primary dressings and topical treatment (15%) to surgical debridement (37.6%), amputation (62.4%), and femoropopliteal revascularization (4.6%). More severe ulcers, classified as grades 4 and 5 on the Wagner scale, were more frequently associated with amputations. By the end of the hospitalization period for these 109 patients, the total hospital expenses exceeded 486,000 reais, averaging R$ 4,461.04 ± 2,995.30 per patient [10].

The highest costs were related to hospital expenses (57%), followed by medical fees (23%), medications (17%), and complementary exams (3%), corroborating the results of this study, which highlight the high costs of hospital expenses. However, the same study indicates that SUS funding was seven times lower than the total expenditure, revealing a discrepancy in the reimbursement values. Furthermore, it was observed that patients who underwent primary healing and minor amputations required longer hospitalization periods, increasing hospital costs [10].

It is estimated that SUS performed 102,056 amputation surgeries, with 70% resulting from complications of Diabetes Mellitus, significantly impacting the financing of public healthcare in Brazil [11]. This information aligns with data from other countries, such as the Netherlands and Spain, where hospitalization periods were longer, but amputation rates were lower [12,13].

From this perspective, it is necessary to assess not only the economic impact but also the social and human costs. Amputations are considered a significant public health issue as they lead to disabilities, social and psychological distress, lifestyle changes, financial difficulties for patients, prolonged hospital stays, and increased mortality rates [14–16].

Studies indicate that amputations bring radical changes to patients’ daily lives, affecting their functional abilities, autonomy, and physical dependence for daily activities. This can lead to an exacerbated emotional burden, with significant psychological impacts, including depression, anxiety, fear, grief over the lost limb, and social discomfort [16].

In this context, the Unified Health System (SUS), recognized as one of the largest and most complex healthcare systems in the world, plays a crucial role in promoting and providing healthcare services to the population. It extends beyond medical care and hospital-based models, actively incorporating primary healthcare actions, including medium and high complexity hospital services, sanitary and epidemiological surveillance, environmental monitoring, and pharmaceutical assistance, which also includes the distribution of chronic-use medications [17].

All these services are part of a set of public policies integrated into SUS, aiming to reduce inequality based on its constitutional principles of universality, equity, and comprehensiveness. These principles contribute to the country’s economic and social development, as healthcare directly impacts quality of life and population well-being. The funding of this complex system is a public responsibility, structured in a tripartite manner, involving contributions from the federal, state, and municipal governments, as well as taxpayer contributions [17].

As a major public health concern, uncontrolled diabetes is among the conditions that generate the highest expenses for SUS, particularly as it is one of the leading causes of non-traumatic amputations, which require prolonged hospitalization and consequently incur higher costs to cover patient expenses during recovery [18].

With the establishment of the Family Health Program (PSF) in Brazil, improvements and advancements in primary healthcare have enabled the development of structured healthcare actions through multidisciplinary teams. This contributes to the planning and implementation of health education and disease prevention strategies [17].

Thus, primary care serves as the foundation for the entire healthcare service flow. Strengthening preventive actions at this level helps avoid severe complications, particularly NCDs, which lead to costly and complex hospital care. Many of these conditions could be prevented or even avoided if health education initiatives were effectively disseminated and easily accessible to all citizens [17].

Among the numerous initiatives within SUS, the structuring of pharmaceutical assistance stands out as it ensures access to essential medications for low-income populations. Medication expenses constitute a significant portion of family health expenditures. Public spending on medications within the free access policy continues to rise and represents the second-largest healthcare expenditure, surpassed only by hospital care. The high prevalence of chronic diseases, particularly among the most vulnerable populations, further increases medication consumption, highlighting the need for greater financial investment [19,20].

Other strategies reinforce the care continuum for NCDs, such as Hiperdia, a program within the Family Health Strategy (ESF) aimed at monitoring and educating patients with hypertension and diabetes. This program seeks to promote health education tailored to patients’ conditions while monitoring the risks of disease-related complications [21].

In this strategy, a multidisciplinary approach is adopted, with professionals within their respective fields setting goals and actions for health promotion and complication prevention. Patient registration facilitates long-term follow-up and generates essential data for healthcare management bodies to allocate further investments in this sector. Within the context of primary healthcare investments, the Previne Brasil program was established through Ordinance No. 2,979, dated November 12, 2019 [21].

This new funding model modifies the distribution of financial transfers to municipalities based on three criteria: weighted capitation, performance-based payment, and incentives for strategic actions. It represents a new financing proposal for primary healthcare, primarily focused on increasing public access to primary care services, thereby expanding patient interaction with healthcare teams. However, there is no guarantee of rapid and effective care, as this would require increasing healthcare teams to support the growing assisted population, representing yet another challenge in the efficient management of NCDs [22].

Despite current strategies for diabetes management, such as the provision of insulin and regular medical follow-ups, Brazil faces challenges in ensuring comprehensive care for people with Diabetes Mellitus, mainly due to socioeconomic disparities. The lack of universal access to adequate treatment, coupled with resource shortages in some regions, makes disease management a complex challenge. Many diabetes patients do not receive the necessary follow-up, leading to severe complications, increased hospitalizations, overburdening the healthcare system, and escalating costs [23].

One limitation of this study is the possibility of underreporting in the data collection and recording system. This may occur due to communication failures or incomplete information, compromising data accuracy and comprehensiveness. Another limitation is the delay in updating the data used in the study, as updates do not occur continuously or immediately. This discrepancy may affect the temporal relevance of findings, especially in the analysis of dynamic or rapidly changing phenomena.

## Conclusions

The study in question highlights, through its results and supporting literature, that Diabetes Mellitus (DM) imposes substantial costs on the public healthcare system in Brazil, with hospital expenses being the most significant. It is also observed that the majority of hospitalization expenditures are allocated to the emergency care network. Regarding the demographic profile of the treated population, there is currently a predominance of male patients and individuals of mixed ethnicity. The study further evidences a shift in patient demographics over the analyzed years, as the previously dominant profile consisted of female and white individuals.

From this perspective, it is understood that DM necessitates significant investments in continuous treatments, medications, and specialized healthcare services. Beyond the financial burden, the societal and individual impacts are considerable, as the disease compromises quality of life, increases workplace absenteeism, reduces productivity, and may lead to physical disabilities and psychological disorders. These factors contribute to intangible costs that further exacerbate the economic burden associated with the disease.

Given this scenario, it is imperative to invest in preventive measures and early diabetes management. The promotion of healthy habits, health education, early screening, and the strengthening of primary health care are essential strategies to prevent disease progression and, consequently, the need for hospitalizations. Investing in these actions can significantly reduce the incidence of severe complications, alleviate hospital overburden, lower hospitalization-related costs, and enhance the quality of life for individuals with diabetes.

## Data Availability

All data files are available from the DATASUS database (link: https://datasus.saude.gov.br/informacoes-de-saude-tabnet/).

## Declarations

## Ethics approval and consent to participate

Not applicable.

## Consent for publication

Not applicable.

## Availability of data and materials

The data were extracted from the Hospital Information System of SUS (SIH/SUS), from the DATASUS platform, which is managed by the Brazilian Ministry of Health and has open access on the website: https://datasus.saude.gov.br/informacoes-de-saude-tabnet/.

## Competing interests

The authors declare that no competing interests exist regarding this study.

## Funding

This study will be partially funded by the Coordination for the Improvement of Higher Education Personnel (CAPES): Financial Code 001. The funders had no role in the study design, data collection and analysis, decision to publish, or preparation of the manuscript.

## Author’ contributions

LLA, AEOM contributed to the conception and design of the work; LLA, KTSC, MESN, KSM, RARS to the acquisition, analysis and interpretation of data; TTXN, MASO, CNS, ACRS, AEOM wrote the work or substantially revised it.

## Acknowledgements

We would like to thank the Federal University of Rio Grande do Norte and the Coordination for the Improvement of Higher Education Personnel (CAPES).

## List of abbreviations

AIH: Hospitalization Authorization
CEP: Research Ethics Committee
DATASUS: Department of Informatics of the Unified Health System
DM: Diabetes Mellitus
ESF: Family Health Strategy
GDP: Gross Domestic Product
IPEA: Institute for Applied Economic Research
NCD: Non-Communicable Diseases
PSF: Family Health Program
SIH: Hospital Information System
SUS: Unified Health System

## Notes

### Competing Interest Statement

The authors have declared no competing interest.

### Funding Statement

The author(s) received no specific funding for this work.

